# A Simple, SIR-like but Individual-Based *l-i AIR* Model: Application in Comparison of COVID-19 in New York City and Wuhan

**DOI:** 10.1101/2020.05.28.20115121

**Authors:** Xiaoping Liu

**Affiliations:** Department of Medicine, Department of Neuroscience, West Virginia University Health Science Center, Morgantown, WV 26506

## Abstract

COVID-19 has spread around the world with nearly 360,000 deaths from the virus as of today (5/28/2020). Mathematical models have played an important role in many key policy discussions about COVID-19. *SIR* or SIR-derived models are a common modeling technique. However, the application of these models needs to solve complicated differential equations, enabling use of these models only by professional researchers. In this study, a simple, SIR-like but individual-based model, the *l-i AIR* model, is presented. The parameters *l* and *i* represent the length of the latent period and the infectious period, respectively. The variable A stands for the number of the infected people in the active infectious period, I for the number of cumulative infected people, and *R* for the number of the people in recovery or death. The nth terms of the three variables are derived, which can be easily calculated in Microsoft Excel, making the program easy to be used in most offices. A transmission coefficient *k* and a transient incidence rate α of the infected people are induced in the model to examine the effect of social distancing and the testing capacity of coronavirus on the epidemic curves. The simulated daily new cases from this *l-i AIR* model can fit very well with the reported daily new cases of COVID-19 in Wuhan, China and in New York City, USA, providing important information about latent period, infectious period and lockdown efficiency, and calculating the number of actual infected people who are positive in antibodies.

## Introduction

In early December 2019, a new type of pneumonia, which is named COVID-19 by the World Health Organization (WHO), was found in Wuhan, Hubei province, China [1, 2], and then the disease was found in all provinces of China and almost all countries around the world. This extensive spread of disease in the world was officially described as a pandemic on March 11, 2010. Currently (5/28/2020), there are more than 5.8 million confirmed cases, and around 360,000 people are killed by COVID-19 globally. To prevent the disease of further transmission, many cities, states or provinces, and countries declare lockdown. Since the beginning of the COVID-19 epidemic, several papers using *SIR* or SIR-derived models to analyze the characteristic of the COVID-19 have been published[3–7]. The *SIR* model was first published nearly 90 years ago[8]. Here, *S* stands for the number of susceptible, *I* for the number of infectious, and *R* for the number recovered (or immune) individuals[9]. A main assumption to the change of *S* with time, *dS/dt*, is that *dS/dt* is proportional to *S* and *I*. This assumption requires both *S* and *I* be well-mixed rapidly and an individual in *S* and *I* have the same probability appearing at different locations in the whole area studied. The same requirement is needed for people in *R*. So, the *SIR* model can be considered as *a* “well-mixed” based model. However, in actuality, individuals in *S, I* and *R* are not well-mixed in the whole population. Individuals in the real world usually have their own moving path daily. Furthermore, the model needs to solve complicated differential equations to calculate epidemic curves, making the use of the *SIR* model to be limited to professional researchers. In this study, we present a new individual-based *l-i AIR* model, which does not require a well-mixed population. Since we have found simple nth term formulas for all variables in this model, it is easy to implant nth terms of these variables in Microsoft Excel, which can be run in most offices. This new mathematical model is then applied to the analysis of the epidemiological characteristics of infectious diseases; and we compare COVID-19 in Wuhan, China and in New York City (NYC), USA, where the lockdown intervention was applied to slow down the disease transmission during the epidemic outbreak.

## Theory: *l-i AIR* Model

If *a* person is infected by an infectious disease, the person will experience *a* latent period (l), an infectious period (*i*), and then recovery or death. In this model, *l* and *i* are two parameters; and the three letters *A*, *I* and *R* are variables. *A* stands for the active infectious individuals or the individuals in the infectious period, *I* for the cumulative infected individuals, and *R* for the recovered individuals. It is assumed that (a) the length of latent period is *l* days or *l* time units (1 time unit can be 1 day or less/more than 1 day); (b) the length of infectious period is *i* days or *i* time units; and (c) the infectious individual infects one person per day or per time unit in the infectious period. Under these assumptions, if *i* = n, it means that an infectious individual will infect 1 person per day (or per time unit) for n days (or n time units) in the infectious period, or will infect a total of n people in the infectious period before the infectious individual is recovered or dead.

In the following, a special example is given for examining how an infectious person spreads the diseases in the *l-i AIR* model. At first, it is assumed that the time unit is 1 day/unit; the latent period is 2 days (*l* = 2); the infectious period is 3 days (*i* = 3); and the first infected individual has been generated on day 1. A column of 5 cells (Figure 1A) is used to represent the infectious status of an infected individual. The transmission pattern of the epidemics is illustrated in Figure 1B:

**Figure 1.**
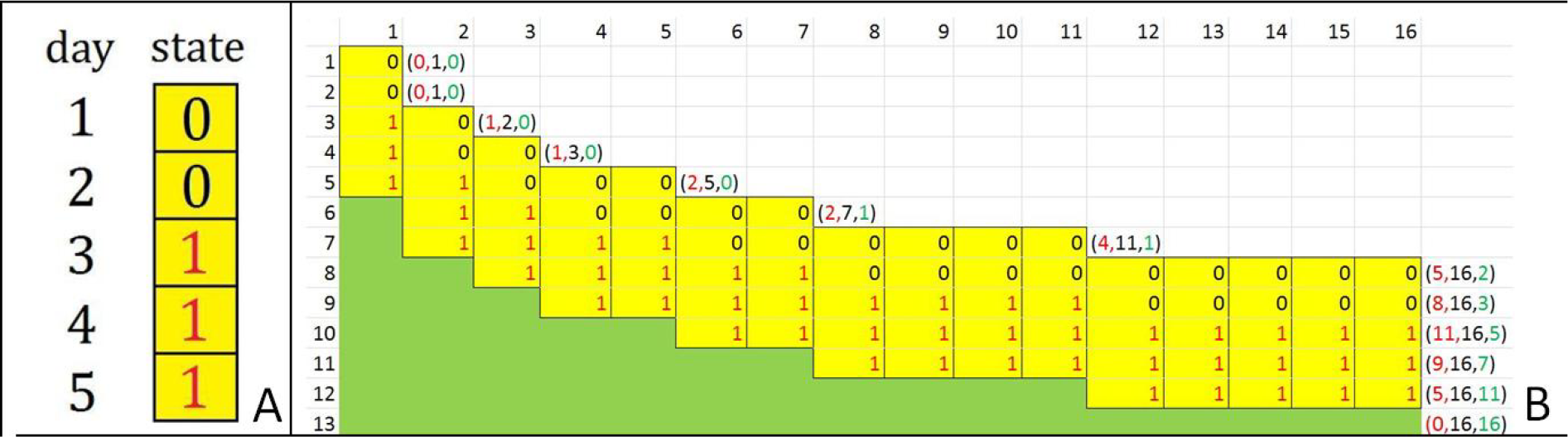
Illustration of *l-i AIR* model. (A) A single column with 5 cells is used to represent the infectious state of an infected individual in the 5 days (*l* = 2 days and *i*=3 days). At day 1 and 2, the state number is 0 meaning that the oerson is in the latent state. From day 3 to day 5, the state

On day 1, the infected individual is in the latent period, so *A*_1_ = 0 because there is no one in the infectious period. However *I*_1_ = 1 because the first infected individual exists already. There is no one recovered, so *R*_1_ = 0. Thus, the three variables (*A*_1_, *I*_1_, *R*_1_) = (0,1,0).

On day 2, the first infected individual is still in the latent period, who is not able to infect others and is not recovered yet, so (*A*_2_,*I*_2_, *R*_2_) = (0,1,0).

On day 3, the first infected person enters the infectious period, so its infectious status changes from 0 to 1 (*A*_3_ = 1). In this infectious status, the individual can infect one person (*I*_3_ = *I*_2_+1 = 2). The new infected person is in the latent period. No one is recovered (R_3_ = 0).Thus, we have (*A*_3_, *I*_3_, R_3_) = (1,2,0).

On day 4, the first infected person is in the infectious period; the second infected person is still in the latent period; so there is only one person in the infectious period (A_4_ = l), and they can only infect 1 more person (*I*_4_ = *I*_3_+1 = 2+1 = 3), and there is still no recovered persons (R_4_ = 0). So, we have (*A*_4_, *I*_4_, *R*_4_) = (1, 3, 0).

On day 5, in addition to the first infected person, the second infected person also enters the infectious period (*A*_5_ = *A*_4_+1 = 1+1 = 2). Since *A*_5_ = 2, they can infect two more persons, or (I_5_ = *I*_4_+2 = 3+2 = 5). There are no recovered people (*R*_5_ = 0). Thus we have (*A*_5_, *I*_5_, *R*_5_) = (2,5,0).

On day 6, the first infected person is recovered, so we have *R*_6_ = *R*_5_+1 = 1. The number of persons in the infectious period decreases by 1, but the third infected person enters the infectious period, so we have A_6_ = *A*_5_-1+1 = 2. Since *A_6_=2*, they can only infect two more persons. Thus *I*_6_ = *I*_5_+2 = 5+2 = 7, or (*A*_6_, *I*_6_, *R*_6_) = (2,7,1).

On day 7, there is still only the first infected person in the recovered status (R_7_ = 1). In addition to infected persons 2 and 3, infected persons 4 and 5 enter the infectious period (*A*_7_ = *A*_6_+2 = 4). Infected persons 6 and 7 are in latent period, so they can only infect 4 more persons, *I*_7_ = *I*_6_+4 = 11. Thus (*A*_7_, *I*_7_, *R*_7_) = (4,11,1).

On day 8, in addition to the first infected person, the second infected person is also recovered (*R*_8_ = *R*_7_+1 = 2). Thus, the number of the infectious persons needs to be subtracted by 1. However, the infected persons 6 and 7 enter the infectious period, so the number of the total infectious persons A_8_ = *A*_7_-1+2 = 5. Since *A*_8_ = 5, they can infect 5 more persons, so *I*_8_ = *I*_7_+5 = 16. Thus, (*A*_8_, *I*_8_, *R*_8_) = (5,16,2).

Considering all of the above steps, we obtain the following formulas to calculate the nth term (*A*_n_,*I*_n_,*R*_n_) after its previous terms are determined. Given *l* = 2 and *i* = 3, we let the first term (*A*_1_,*I*_1_,*R*_1_) = (0,1,0) and all previous terms as n<1 are (0,0,0), then we have the following formulas for calculating the nth term:

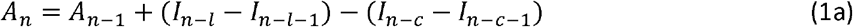

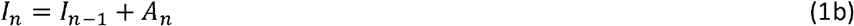

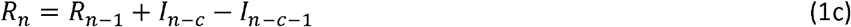

In the above equations, *c* = *l+i*. From Eqn. (1) we can easily find the total infected person in the latent period and infectious period:

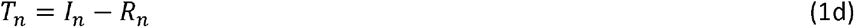

In the above example, we have *l* = 2 and *i* = 3, so we can calculate the second term from the first term (0,1,0) considering that *A*_m_ = 0 and *R*_m_ = 0 as *m*≤1, *I*_1_ = 1 at m = 1 and *I*_m_ = 0 as m<1.

On day 2:

*A*_2_ = *A*_1_ + *I*_0_-*I*_−1_ – (*I*_−3_ – *I*_−4_) = 0 + 0 − 0 − (0 – 0) = 0

*I*_2_ = *I*_1_ + *A*_2_ = 1 + 0 = 1

*R*_2_ = *R*_1_ + *I*_−3_ — *I*_−4_ = 0 + 0 — 0 = 0

Thus, (*A*_2_, *I*_2_, *R*_2_) = (0,1,0)

In the same way, we can use Eqn. (1) to calculate (*A*_n_, *I*_n_, *R*_n_) from n = 3 to n = 8. The calculated results are exactly the same as these (*A*_n_, *I*_n_, *R*_n_) terms derived from the transmission pattern showing Figure 1B.

## *l-i AIR* model with lockdown intervention

If the outbreak of an epidemic occurs in a geographic area, and the area is lockdown from a day (n = m) to slow down or stop the epidemic outbreak, then we can simply revise Eqn. (1b) from the lockdown day (n = m):

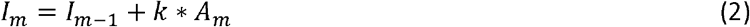

Where the transmission coefficient *k*=0 if assuming that the lockdown intervention completely blocks any further infection to uninfected persons, or *k* is a positive value close to zero indicating that the transmission activity has been greatly reduced. To demonstrate the effect of lockdown on *A*_n_, *I*_n_ and *R*_n_, we assume that the lockdown starts at day 9 in the above example:

On day 9, one infected person (the third infected one) is recovered, but 4 infected persons enter the infectious status from latent status. Thus, the number of the recovered people increases by 1, or R_9_ = *R*_8_+1 = 3; and the number of the active infectious persons A_9_ = *A*_8_-1+4 = 8. Since one infected person is recovered and no more persons are infected because of the transmission coefficient k = 0 after lockdown intervention, we have *I*_9_ = *I*_8_+k*A_9_ = *I*_8_ = 16. Thus (*A*_9_, *I*_9_, *R*_9_) = (8,16,3).

On day 10, two more persons are recovered (*R*_10_ = *R*_9_+2 = 5), but 5 new persons enter the active infectious period (*A*_10_ = *A*_9_-2+5 = 8–2+5 = 11). Since no new persons are infected because of k = 0, we have *I*_10_ = *I*_9_ = 16, so (*A*_10_,*I*_10_,*R*_10_=(11,16,5).

On day 11, two persons are recovered (*R*_11_ = *R*_10_+2 = 7), but no new active infectious persons are added, so we have *A*_11_ = *A*_10_–2 = 11–2 = 9 and *I*_11_—*I*_10_ = 16, thus (*A*_11_, *I*_11_, *R*_11_)=(9, 16, 7).

Similarly, on day 12 and 13, 4 persons and 5 persons are recovered, respectively. So, we can get (*A*_12_;*I*_12_,*R*_12_)=(5,16,11) and (*A*_13_,*I*_13_,*R*_13_)=(0,16,16).

The above (*A*_n_, *I*_n_, *R*_n_) terms were derived from the transmission pattern shown in Figure 1B. These terms can be also calculated from Eqn. (1) & (2) assuming that *k* = 0 after lockdown intervention. If *k*=0, the nth term for *I*_n_ can be simplified as

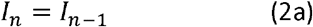

Thus we can calculate *A*_n_, *I*_n_, *R*_n_ on day 9:

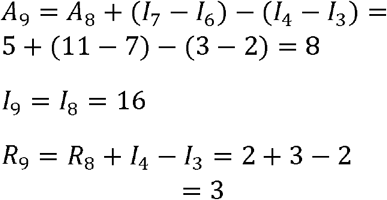

So, we have (*R*_9_, *I*_9_, *T*_9_) = (8,16,3). Similarly, we calculate other terms from n = 10 to n = 13. These calculated terms are exactly the same as those we have demonstrated above.

## Simulated results

To simulate an epidemic outbreak, *l* and *i* can be given different numbers for fitting the reported data. Eqns. (1) and (2) can be easily implanted into Microsoft Excel to examine how *A*_n_, *I*_n_ and *R*_n_ change with n. Here, n can be considered as a unit of time or date. By the following simulation, we can examine the characteristics of *A*_n_, *I*_n_ and *R*_n_ in the outbreak phase and lockdown phase.

1. No epidemic outbreak at *i* = 1. The rapidity of outbreak of epidemics is greatly related to the time length of *l* and *i*. In Figure 2, we demonstrate how the total number of infected individuals (*T*_n_) changes with time (n) for different *l* at *i* = 1. No epidemic outbreak can be seen from the figure. At *i*=1, the present infectious person will only infect one new person before the former person loses his/her transmissibility by recovery. As a result, the number of infectious individuals can’t be accumulated up to more than 2. Although the total number of infected persons in the latent and infectious periods (*T*_n_) jumps up and down between 1 and 2 repeatedly, the number of recovered individuals (*R*_n_) can be accumulated up with time. However, the number of infectious persons (*A*_n_), similar to *T*_n_, jumps between 0 and 1 repeatedly (Figure 3).

**Figure 2.**
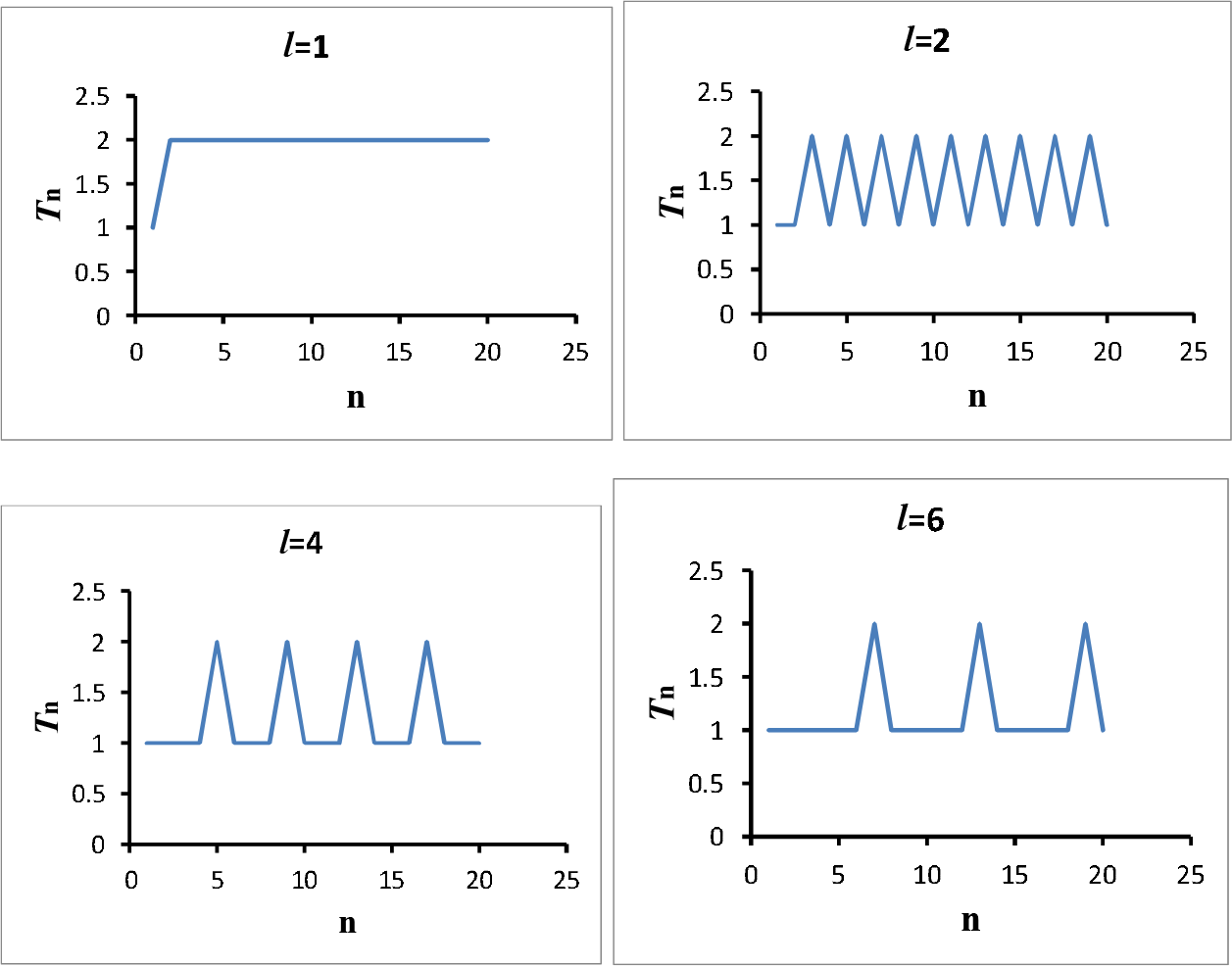
Changes of the total number of infected individuals in both the latent and the infectious periods *T*_n_ with n for different *I* at *i* = 1. The values of *T*_n_ vary between 1 and 2.

**Figure 3.**
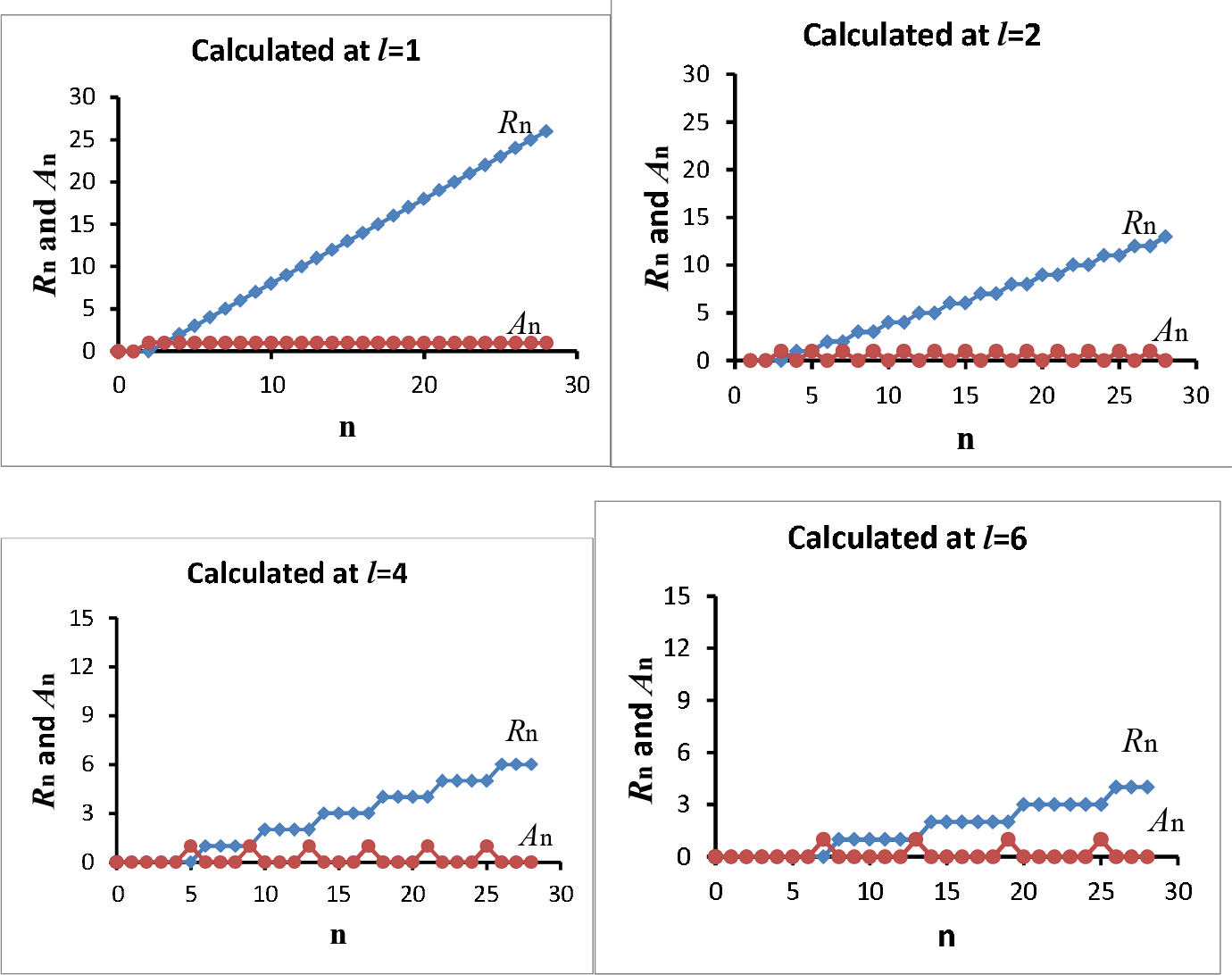
Changes of *R*_n_ and *A*_n_ with time (n) for different values of *I* at *i* = 1.

2. Propagated epidemic curves as *l*>*i*>1. Propagated epidemic curves usually consist of a series of waves with successively larger peaks[10]. By simulations, we observed that the propagated epidemic curves are formed when *l*>*i*>1. In Figure 4, we illustrate three calculated curves of *A*_n_ for *l* = 4, 6 and 8 at *i*=2. These calculated curves are very similar to the propagated epidemic curves reported in the literature.

**Figure 4.**
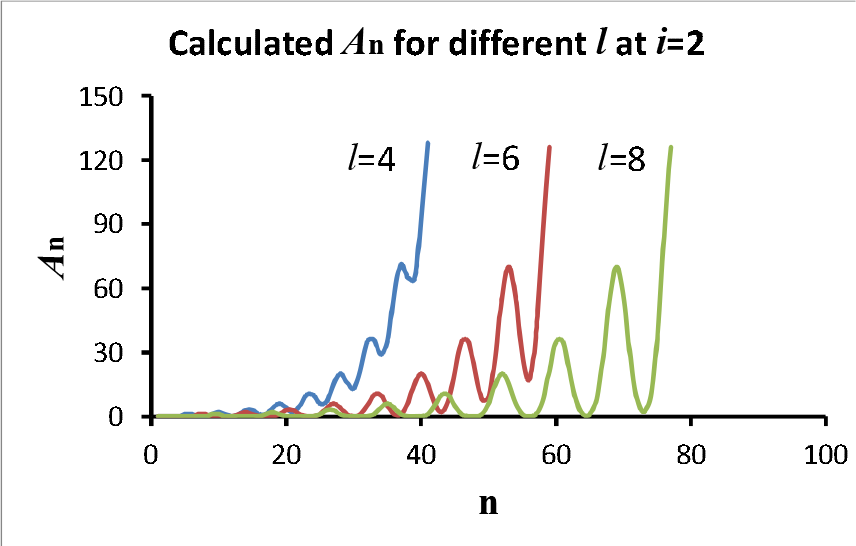
Propagated epidemic curves for *I*=4, 6 and 8 at *i*=2. Each curve consists of a series of waves with successively larger peaks.

3. Exponential epidemic outbreak as *i*≥*l*≥1 except *i*=1. In the early stage of epidemic outbreak, it is often seen that epidemic curves of new cases exponentially grow with time. By simulation using the *l-i AIR* model, we can see that all curves of *A*_n_, *I*_n_ and *R*_n_ exponentially increase with time if *i*≥*l*≥1 except *i*=1 except *i*=1. A special example of the calculated *A*_n_, *I*_n_ and *R*_n_ at *I* = 2 and *i* = 4 is shown in Figure 5A. The logarithm of *A*_n_, *I*_n_ and *R*_n_ at and *i* = 4 is linear with n (Figure 5B), indicating that *R*_n_, *I*_n_ and *T*_n_ exponentially increase with n.

**Figure 5.**
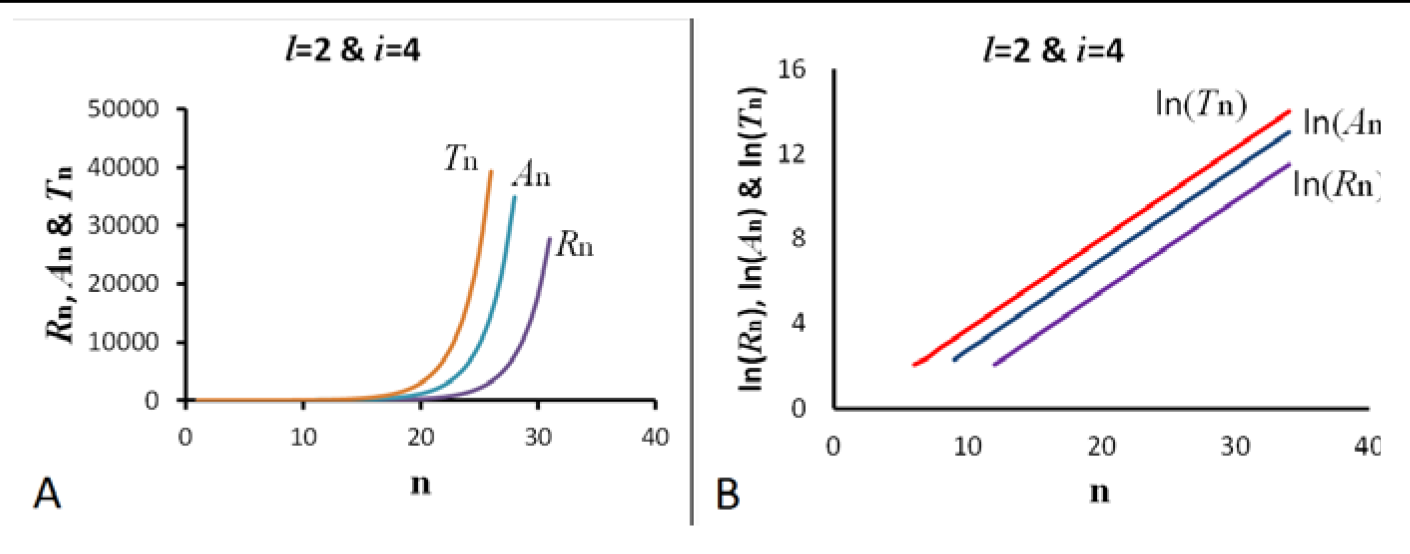
An example of exponential growth of *R*_n_, *A*_n_ and T_n_ as *i*>*I*>1. (A) The curves are calculated from Eqn. 1 assuming *I* = 2 and *i*=4. (B) The logarithm of *A*_n_, *I*_n_ and *R*_n_ linearly increase with n.

4. Epidemic curves with lockdown intervention. During an epidemic outbreak, if the lockdown intervention of the epidemic area is performed, the forward tendency of the epidemic curve will change depending on how strict the lockdown intervention is being implemented. In an ideal situation, the infection of an infectious person to an uninfected person is completely blocked, or the transmission coefficient *k* in Eqn. (2) is 0, then Eqn. (2) can be simply replaced by Eqn. (2a). Using Eqn. (1) and Eqn. (2a), we calculated *R*_n_, *A*_n_ and *T*_n_ at different combination of *l* and *i*. The calculated *A*_n_ and *T*_n_, assuming that the first infected person appears on day 1 and that the lockdown is performed on day 41, are demonstrated in Figure 6.

**Figure 6.**
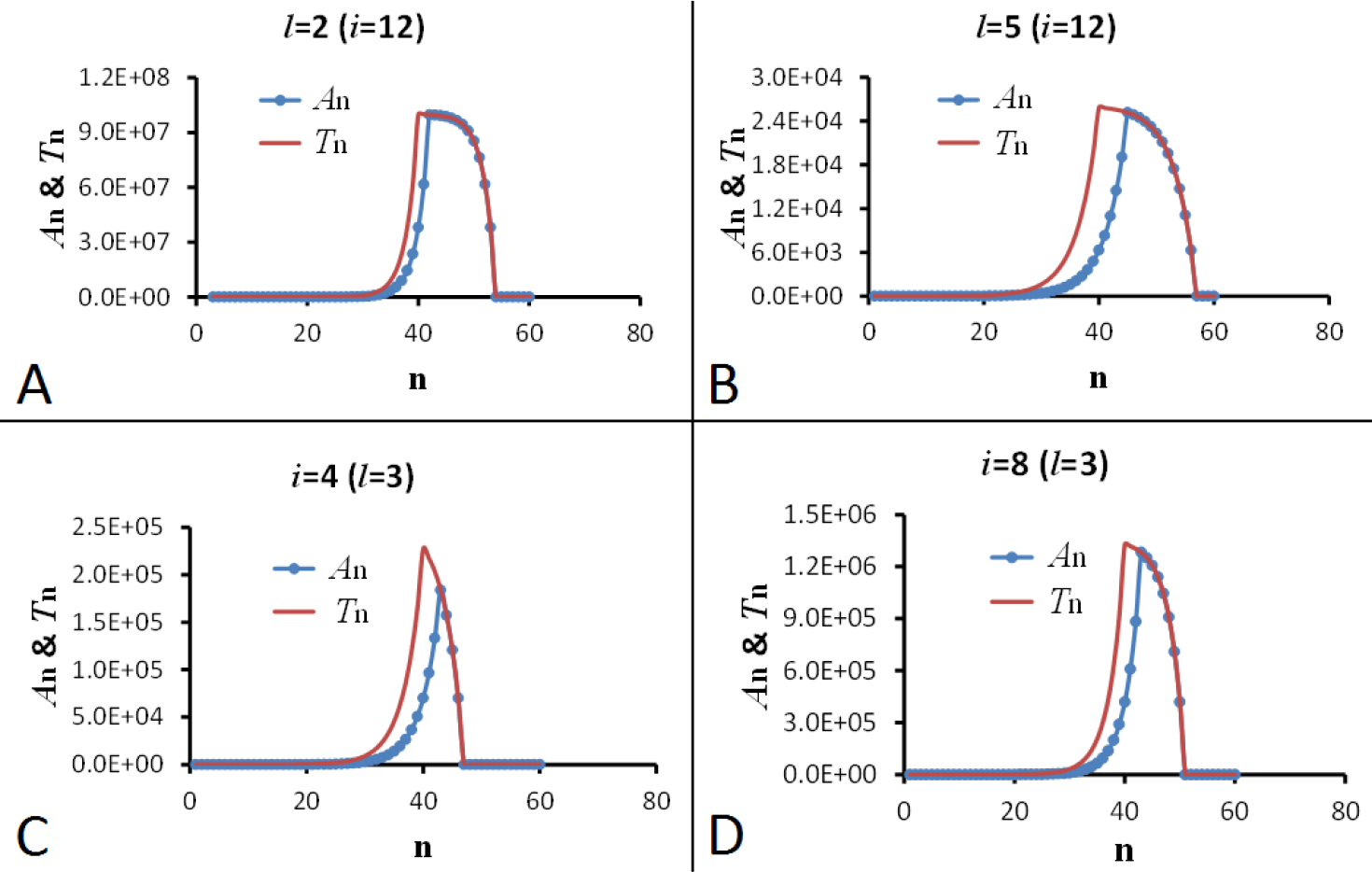
Calculated epidemic curves (*A*_n_ & *T_n_)* at 4 different combinations of *I* and *i* assuming the lockdown intervention started at n = 41 and the transmission coefficient *k* = 0 after lockdown.

In comparison, if the virus transmission is not completely blocked by the lockdown intervention or *k* is greater than 0, then the shape of epidemic curves will depend on *k*. In Figure 7, we present the simulated epidemic curves for the nonzero value of *k* at *l* = 3 and *i* = 5 to show how the shape of *A*_n_ curve changes with *k* (*A*_n_ is proportional to the number of the reported daily new cases).

**Figure 7.**
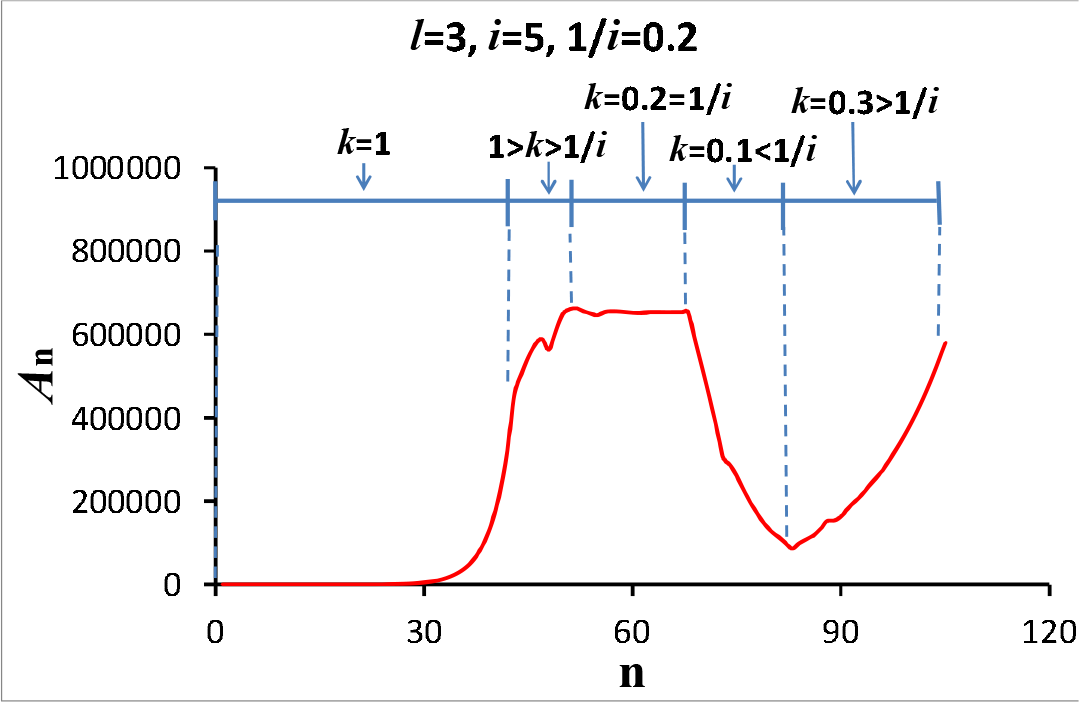
Simulated epidemic curves with lockdown intervention for different transmission coefficient *k*.

When the number of the cumulative infected individuals increases to a range comparable to the total susceptible population, the remaining susceptible people is largely reduced. In this case, the chance to contact an uninfected person is reduced, so *k* will decrease even If there is no lockdown intervention. Assuming that *k* is proportional to the ratio of the number of uninfected people (*N-I*) to the number of initial susceptible population (*N*), *k*_n_=(*N-I*_n-1_)/*N*, the simulated epidemic curves of *A*_n_, *I*_n_ and *R*_n_ at *l* = 1, *i* = 2 and *N* = 1×10^6^ are demonstrated in Figure 8. As defined previously, *I* is the number of the cumulative infected people, and (*N-I*) is the number of the total uninfected people. During the epidemic outbreak, *I* increases from 0 to a number close to *N* so *k* will decrease from 1 to a number close to 0 correspondingly.

**Figure 8.**
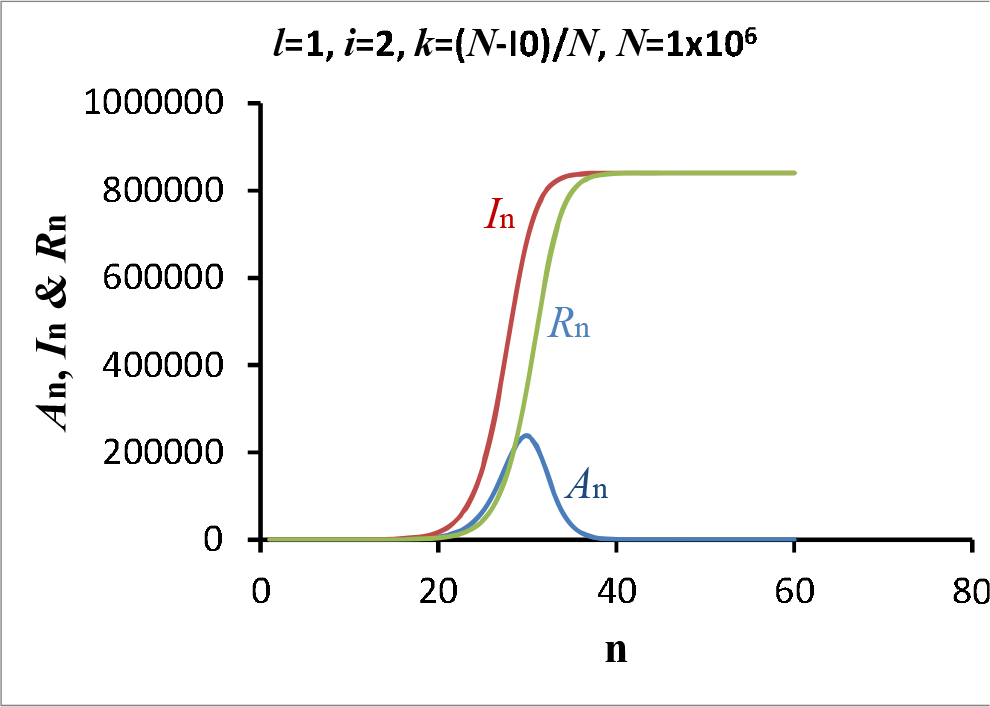
The simulated epidemic curve of *I*_n_ assuming the susceptible population *N* is 1 million.

## Application of the *l-i AIR* model to compare COVID-19 outbreak in Wuhan, China and in NYC, USA

### Simulation of COVID-19 outbreak in Wuhan.

In the *l-i AIR* model, *A* stands for the number of individuals in the infectious period. To compare the calculated epidemic curve with the reported data in an epidemic event, we need to have an assumption: the daily new case (*y*) of the epidemic event can be considered as the transient incidence of infectious disease at day n, which is proportional to *A*_n_, or *y* = *aA*_n_.

Here *a* is the incidence rate. In Figure 9A, we compare the calculated daily cases with the reported daily new cases of COVID-19 in Wuhan, China. The red line with closed circles represents the three-day averages of the reported daily new cases in Wuhan. The daily new cases were reported by Hubei Health Commission. Between 2/12/2020 and 2/18/2020, a total of 7 days, clinical diagnosis of COVID-19 was added as a diagnostic criteria, resulting in large increase in case number on February 12 and subsequent days. This change in diagnosis of COVID-19 disease creates difficulties in using the reported cases for data analysis[4]. Fortunately, both the number of the total new cases and the number of cases diagnosed clinically were listed in the first 4 days between 2/12/2020 and 2/15/2020, so it is easy to find the number of daily new cases confirmed from the viral tests by subtracting the clinically diagnosed case number from the total new case number. The daily new cases in the last three days between 2/16/2020 and 2/18/2020 can be estimated from the reported daily new cases by using information (the ratio of the numbers measured by the two diagnosis methods) provided in the previous days. To calculate daily new cases from Eqns. (1) and (2), we set: (a) the first COVID-19 case appeared on December 8, 2019, and the lockdown day of Wuhan started on January 23, 2020; (b) *l* = *l* unit (3 days) [11,12], *i* = 4 units, and each time unit is 3 days; (c) the transmission coefficient *k* is 1 before lockdown; 0.25 in the first time unit (3 days) after the city lockdown considering that each infected individual can still transmit the virus to the person who lives with the infected individual; 0.03 in the second time unit and all 0.02 from the third time unit after the city lockdown; and (d) the transient incidence rate α is 1/100 or 1%. From Figure 9A, it can be seen that the calculated number of daily new cases (green line) is consistent with the reported daily new cases (red line with closed circles) before and on the date 2/12/2020. After this date, it seems that the reported number of daily new cases makes a turn and deviates from the calculated curve. Changing the parameter *k* or *α* can make the calculated curve fit the reported number of daily new cases better. However, although increasing *k* can moderately improve the fitting, a bigger *k* will be too large to be reasonable considering the strict lockdown that was performed during that time. This deviation is more likely caused by changes in α because of some other intervention used during this time, such as the large increase in the number of viral tests and the use of more than 10 Fangcang shelter hospitals to admit more than 10,000 patients[13]. By gradually increasing *α* from 2/15/2020 to 3/1/2020, the calculated number of daily new cases fits with the reported number of daily new cases pretty well (Figure 9B).

**Figure 9.**
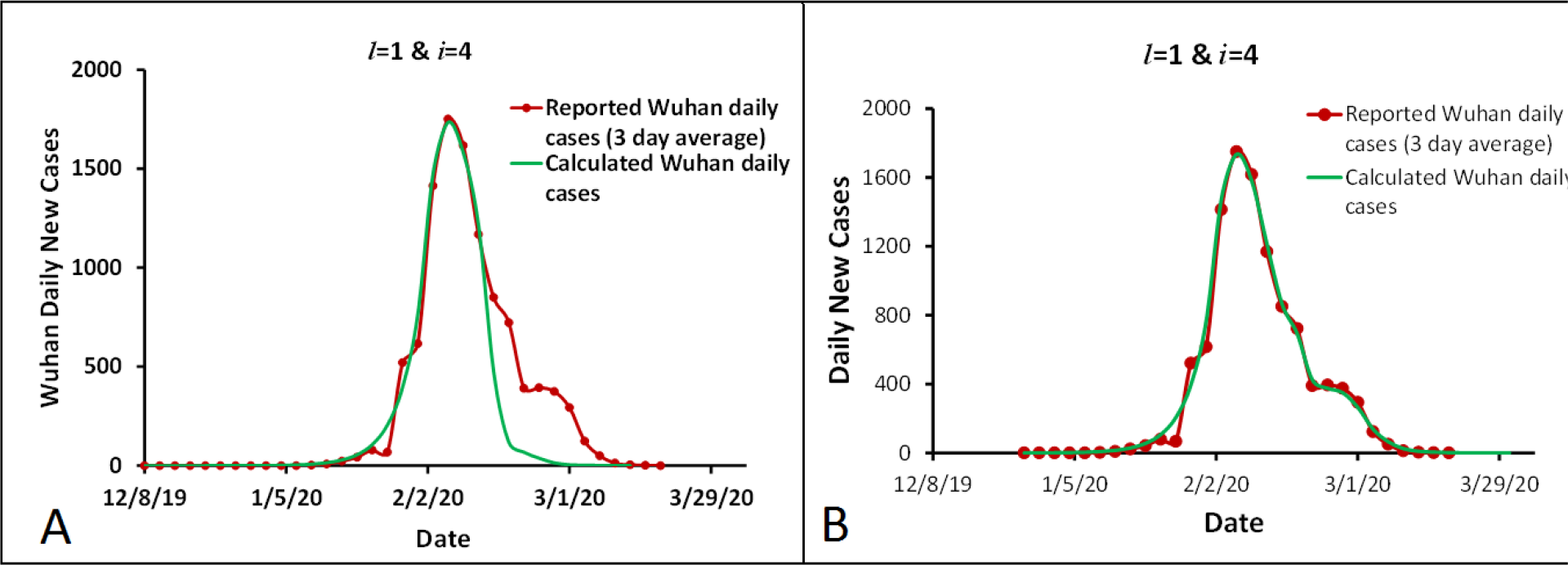
Comparison of the calculated daily new cases with the reported daily new case during the outbreak of COVID-19 in Wuhan.

### Simulation of COVID-19 outbreak in NYC

The first COVID-19 case in New York City was confirmed on March 1, 2020[14], Research indicates that coronavirus started to transmit in the New York state between late January to mid-February[15]. A state of emergency in New York was declared on March 7 after 89 cases were confirmed in the New York state. All schools, bars, and restaurants in NYC were closed on March 17, 2020[16]. The statewide stay-at-home order of New York was effective on March 22, 2020[17], To simulate the reported daily new cases of COVID-19 in NYC, a different combination of *l* and *i*, and different start date of the first infected person who initiated the COVID-19 transmission in NYC were tried. It was found that the simulated curve of daily new cases can fit well with the reported daily new cases if *l* = 4[18], *i* = 10, and the start day of the first infected person appeared in NYC was on February 9, 2020. In the simulation, the transmission coefficient *k* was largely dropped from 1 on March 17 (k = 0.45), the first day that NYC closed schools, restaurants and bars, and then gradually decreased to 0.064 by April 19, 2020. The transient incidence rate α is 1/85 or ∼1.2%. The reported daily new cases in NYC and the simulated epidemic curve are demonstrated in Figure 10A. The calculated number of cumulative infected people (*I*_n_) based on Eqns. (1) and (2) using the parameters for Figure 10A is shown in Figure 10B.

**Figure 10.**
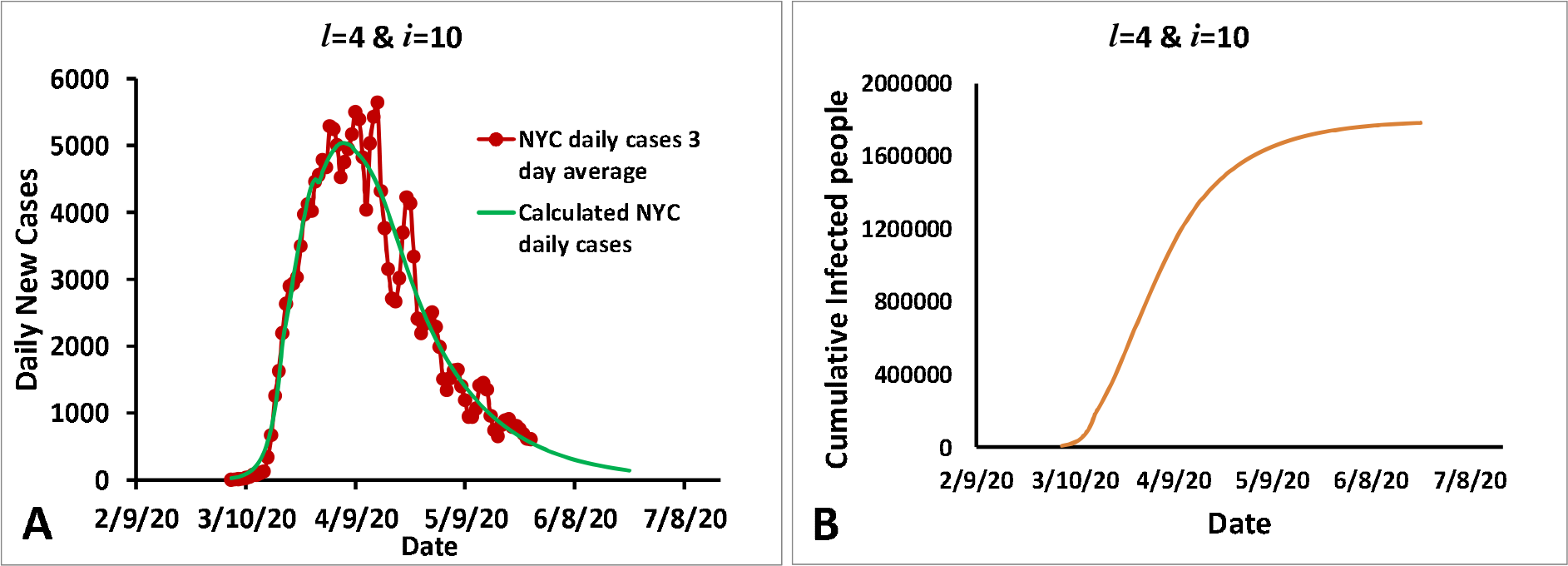
The simulated curve of daily new cases of COVID-19 and the reported daily new cases of New York City assuming that *I* = 4, *I* = 10 and *k* gradually decreases to 0.064 after March 17 (A). The number of cumulative infectious people was calculated from Eons. (1) and (2) (B).

### Comparison of the spreading characteristic of COVID-19 in Wuhan and in New York City

The values of *I* and *i* of COVID-19 in NYC are different from those of the COVID-19 in Wuhan. For Wuhan’s COVID-19, *l* is 1 time unit and *i* is 4 time units, with the time unit being 3 days/unit. This means: (a) the latent period *l* is 3 days; (b) the infectious period *i* is 12 days; and (c) each infectious individual can infect one person every 3 days or infect 4 persons in 12 days. For NYC’s COVID-19, *l* = 4, *i* = 10, and time unit is 1 day/unit. This meansthat the latent period *l* and infectious period *i* are 4 days and 10 days respectively; and each infectious individual can infect 1 person per day for a total of 10 days. To compare the transmission characteristic of the two types of COVID-19, simulations were performed by assuming that the outbreak of COVID-19 occurs in an area having 1 million susceptible people and no intervention is used to slow down the disease outbreak. In the simulations, it was assumed that the transmission efficiency is gradually reduced while the number of uninfected people is gradually reduced with time by assuming that *k* is proportional to (*N-I*)/*N*. The simulated epidemic curves of *A*_n_, *I*_n_ and *R*_n_ are shown in Figure 11. COVID-19 in Wuhan takes 72 days to reach the outbreak peak in an area with 1 million susceptible people, but COVID-19 in NYC only needs 54 days to reach the outbreak peak in the same area with the same population.

**Figure 11.**
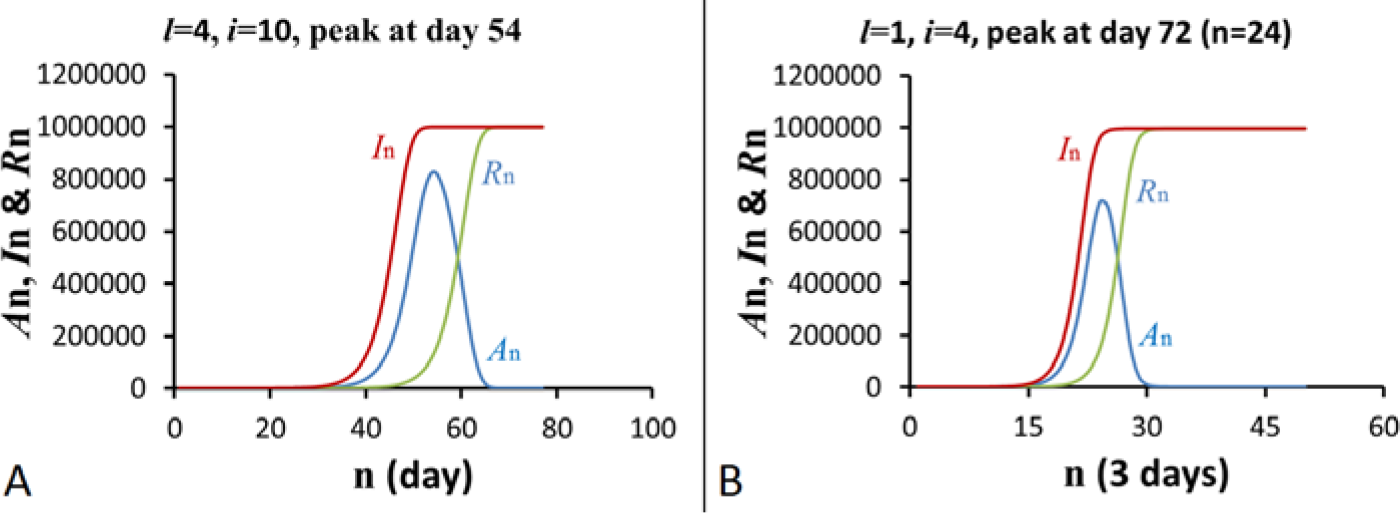
Comparison of transmission characteristic of COVID-19 in New York City, USA (A) with that in Wuhan, China (B).

## Discussion

Simulations based on Eqn. (1) (Figure 2 & 3) show that no epidemic outbreak can happen if *i* = *1* regardless of the length *l* of the latent period, even though the number of the total recovered people (*R*_n_) will be linearly accumulated up with time. The meaning of *i* = *1* is that an infected individual can only pass the virus to one another person before the infected individual is recovered or dead. Under this situation, *T*_n_ and *A*_n_ cannot be accumulated with time after the transmission process of the disease started by the first infectious individual, so this transmission process may be stopped any time when an infectious individual is not able to infect another person before recovery because of any interference.

For any other situations of *i*>1, all of *A*_n_, *T*_n_ and *R*_n_ will continuously increase with time until an intervention is applied or the number of available susceptible people has been largely reduced. For special conditions of *l*>*i*>1, simulations (Figure 4) show the repeated wave-shape in the epidemic curves, which have been observed in the real world, named as propagated epidemic curves, usually transmission by direct person-to-person contact[10]. From the viewpoint of the *l-i AIR* model, the propagated epidemic curves can happen when *l*>*i*>1.

For all other conditions of *i*≥*l*≥1 and *i*>1 at *l* = 1, all of *A*_n_, *T*_n_ and *R*_n_ will exponentially increase with time until an intervention is applied or the number of available susceptible people is largely reduced (Figure 5). As shown in Figure 6, if the transmission process is completely blocked by a lockdown intervention or *k*=0, then *T*_n_ will stop to increase on the lockdown day and decrease to 0 in *l*+*i* days (the total length of the latent period and the infectious period). In comparison, An will stop to increase *l* days later, but will decrease to 0 in *i* days. As a result, both *T*_n_ and *A*_n_ decrease to 0 on the same day even though *A*_n_ stops to increase or reaches the peak *l* days later. Since *A*_n_ is proportional to the reported daily new cases in an epidemic event, the effect of lockdown on the daily new cases will be observed *l* days later. This is reasonable because those individuals infected immediately before the lockdown starting day will get symptoms after the latent period of *l* days (assuming that time in latent period and time in incubation period are similar to each other), so the daily new cases will continuously increase for *l* days after the beginning of the lockdown intervention. Figure 6A & 6B also show that for a given *i* = 12, increasing *l* from 2 to 5 reduces the peaks of the epidemic curves by nearly 4 orders of magnitude. In contrast, for a given *l* = 3, increasing *i* from 4 to 8 only increases the peak by 6 folds.

If the lockdown process is not strict enough in such a way that the transmission process is not completely blocked, then *k* will be greater than 0. In this situation, the epidemic curve may decrease, go flat or even continuously increase depending on whether *ki* is less than 1, equals 1, or greater than 1. Here, *ki* is similar to the basic production number R_0_ used in the SIR model[19]. A simulation is demonstrated in Figure 7, assuming that *l* = 3 and *i* = 5. When *ki* = 1, *k* will equal 1/*i*=0.2. It can been seen that the epidemic curve increases when *k*>0.2, goes flat at *k*=0.2, and decreases when k< 0.2. In some situations, the number (*I*) of the cumulative infected individuals becomes large enough to compare with the number of the initial susceptible population *N* so that the chance of the infected individuals to meet uninfected individuals gradually decreases. A simulation was done by assuming that the chance of infected individuals meeting uninfected individuals is proportional to (*N-I*)/*N* (Figure 8). It can be seen that a peak appears in the epidemic curve when *I*_n_ is comparable to *N*. To make the epidemic curve decrease, *I* must be large enough to meet the condition (*N-I*)/*N*<1/*i*. Thus, we have *I*>*N*(1-1/i). If *i* = 2, *I* must be greater than 0.5N to make the epidemic curve decrease. As shown in Figure 8, the peak of *A*_n_ appears after *I*_n_ passes 0.5 *N*.

The COVID-19 pandemic provides a lot of data to test this *l-i AIR* model. The first example is the COVID-19 outbreak in Wuhan, China, where the first COVID-19 patient was seen in early December 2019 [4], After testing different combinations of *l* and *i*, the final simulated epidemic curve (Figure 9) fits the reported daily new cases in Wuhan pretty well by meeting all of the following requirements: (a) the epidemics started on December 8, 2019; (b) the lockdown intervention was started on January 23, 2020; (c) the latent period (*l* = 1 unit or 3 days) is within the range of reported data[11, 12]; and (d) the simulated epidemic curve fits well with Wuhan’s daily new case. It can be noticed that the reported daily cases (the red line with closed circles) are deviated from the simulated epidemic curve (green line) after February 12, 2020. This may be related to interventions implanted during this period, such as the large increase in the number of viral tests and the use of more than 10 Fangcang shelter hospitals to admit more than 12,000 patients between February 5 and March 10, 2020[13]. These interventions can help to find many more patients with mild symptoms, but may generate a greater apparent transient incidence rate of epidemics than usual. By gradually increasing the incidence rate α from February 15 to March 1, 2020, the calculated daily new case fits the reported data very well (Figure 9B). The second example is COVID-19 outbreak in NYC, USA. As shown in Figure 10, the simulated epidemic curve fits well with the reported daily new cases in NYC. The simulated curve meets the following requirements: (a) the first case started on February 9, 2020, which is within the time range estimated in the report[15]; (b) the intervention to slow down the COVID-19 transmission started on March 17, 2020 and more strict interventions were added later; (c) the latent period (*l* = 4 days) is within the range of the reported data; and (d) the simulated epidemic curve fits well with New York City’s daily new cases (Figure 10A). Using the same parameters for Figure 10A, the calculated number *I*_n_ of the cumulative infected people is shown in Figure 10B. On May 1, 2020, *I*_n_ is 1.58 million, which is very close to the estimated 1.67 million people[20] infected with SARS-CoV-2 in NYC that was determined from the results of antibody tests[21].

Research has shown that the earliest cases of COVID-19 in New York were likely brought in by travelers from Europe[15]. Simulated curves in Figure 11 show that COVID-19 in Wuhan takes 72 days to reach the outbreak peak in an area with 1 million susceptible people, while COVID-19 in NYC needs only 54 days to reach the outbreak peak in the same area with the same population. This indicates that the transmission rate of NYC’s COVID-19 is nearly 30% greater than the transmission rate of Wuhan’s COVID-19. Furthermore, the value of *i* for NYC’s COVID-19 is 10, but *i* is 4 for COVID-19 in Wuhan. Because an intervention must reduce *k* to below 1/*i* for making the epidemic curve decrease, it needs to reduce *k* down to 1/10 or 0.1 to make the epidemic curve decrease in New York. However, it just needs to reduce *k* down to 1/4 or 0.25 in Wuhan. Therefore, it is likely that COVID-19 in NYC has stronger infectivity than that in Wuhan.

## Data Availability

All data of daily new cases of COVID-19 were obtained from reliable sources. They are available to public.

https://www.worldometers.info/coronavirus/

https://en.wikipedia.org/wiki/COVID-19_pandemic_in_New_York_(state)

https://www.nytimes.com/2020/05/28/nyregion/coronavirus-ny-live-updates.html

http://www.cnr.cn/hubei/yaowen/20200202/t20200202_524956737.shtml

